# Supplementation of AQUATURM® (Water soluble turmeric extract) prior to and following heavy eccentric exercise alleviates symptoms of Delayed onset muscle soreness (DOMS)

**DOI:** 10.1101/2022.10.08.22280682

**Authors:** Sanjay Tamoli, Narendra Mundhe, Vinay Pawar, Sandip Birari

## Abstract

**Background:** Curcumin, the most active component in turmeric helps attenuate DOMS.

**Objectives:** A clinical study was conducted to evaluate efficacy and safety of AQUATURM® in subjects with Delayed Onset Muscle Soreness (DOMS).

**Methods:** Twenty healthy men in each of the two subsets completed the study. Subjects were assigned as per a computer-generated randomization list. In subset-I, subjects were asked to take given product in a dose of 1 capsule (250 mg) for 2 days. In subset-II, subjects were asked to take given product in a dose of 1 capsule (250mg) for 5 days. In subset-I, eccentric exercise was performed at hour 6. In subset-II eccentric exercise was performed at day 2.5. The outcome measures were muscle pain, tenderness, swelling, jump performance, muscle inflammatory markers, rescue medication and adverse events. These outcome measures were evaluated at 0 hour, at 6 hours (immediately after exercise), at 24 hours and at 48 hours in subset-I, and at day 1, day 2.5 baseline (immediately after exercise), at 24 hours from baseline and at 48 hours from baseline in subset-II.

**Results:** A significant reduction in muscle pain, tenderness and swelling post eccentric exercise was observed in AQUATURM® Capsule group compared to placebo group in both the subsets of the study. Additionally, when compared to the placebo group, the AQUATURM® group showed a statistically significant increase in post exercise average single-leg vertical squat jump. AQUATURM® Capsule was well tolerated by the subjects.

**Conclusion:** Supplementation of AQUATURM® Capsule prior to and following heavy eccentric exercise in healthy men resulted in relieving muscle pain, tenderness and swelling from DOMS.

## Background and Study Rationale

Delayed-Onset Muscle Soreness (DOMS) is classified as a type I muscle strain or pain that occurs due to any unaccustomed activity, especially eccentric exercises. DOMS is most prevalent in athletes when they are returning to training following a period of reduced activity. The symptoms of DOMS include pain, oedema, reduced strength, and range of motion in corresponding joints and muscles.^1,2^ Various mechanisms of action of DOMS have been postulated such as build-up of lactic acid, muscle spasm, connective tissue damage, muscle damage, inflammation and enzyme efflux. ^3,4^

Both nonpharmacological and pharmacological treatments are being practiced for the management of DOMS. Nonpharmacological treatment mainly includes physiotherapy. It is one of the major main stays of the management of DOMS. Other non-pharmacological interventions include cryotherapy, therapeutic ultrasound thermotherapy, stretching, exercises, electrical currents, and soft tissue massage.^5,6,7^ Pharmacological treatment includes non-steroidal anti-inflammatory drugs (NSAIDs) such as diclofenac, aceclofenac, ibuprofen. It has been observed that the analgesic effect of NSAIDs is temporary and dose dependent. Exercise remains the most effective means of alleviating pain during DOMS. ^8, 9^

Several studies have reported that curcumin supplementation attenuates DOMS after exercise.^10,11,12^ Curcumin, the principal curcuminoid and the most active component in turmeric, is a biologically active phytochemical.^13^ Curcumin helps in reducing inflammatory responses that occur during the recovery phase after exercise and thus helps in reducing symptoms of DOMS.^14^

AQUATURM^®^, a proprietary curcumin extract developed by Lodaat Pharma, contains water soluble turmeric extract standardized to not less than 23% total Curcuminoids. Looking at the activities of Curcumin, a hypothesis was postulated that AQUATURM^®^ could be useful in the management of DOMS. To test the hypothesis, a clinical study titled “Evaluation of Efficacy and Safety of AQUATURM^®^ in Subjects Suffering from Delayed onset muscle soreness (DOMS) – A Randomized, Double Blind, Placebo Controlled, Comparative, interventional, Prospective Clinical Study” was conducted.

## Materials & Methods

### Settings and locations

The study was conducted at KVTR Ayurveda College and Hospital, Boradi, Dhule District, India.

### Study design

This was a randomized, double blind, placebo controlled, comparative, interventional, prospective study.

### Ethical considerations

The study was approved by the Institutional Ethics Committee KVTR Ayurveda College and Hospital, Boradi, Dhule District, India and registered with Clinical Trial Registry - India (CTRI/2021/08/035419, Registered on: 04/08/2021]). The study was carried out and reported adhering to CONSORT statement.

### Study duration & Visits

There were two subsets of the study. In the subset-I, the total duration of the study product consumption was 2 days and, in the subset-II it was 5 days. In subset-I, the study visits were planned and outcomes were assessed on day 1 (0 hour), day 1 (6 hour), day 2 (24 hour) and day 3 (48 hour). In subset-II, the study visits were planned and outcomes assessed on day 1, day 2, day 2.5, Day 2.5 (baseline), 24 hours after baseline, and 48 hours after baseline.

### Eligibility criteria for participants

#### Inclusion Criteria

Moderately active (regular aerobic exercise for at least 4 hours per week) healthy male volunteers of age between 21 and 45 years (both years inclusive), with no known musculoskeletal pathology, ready to provide written informed consent and follow the protocol requirements of the clinical study were included in the study.

#### Exclusion Criteria

Subjects with known musculoskeletal pathology, using systemic corticosteroids within 2 months of screening, or using any other investigational product within 1 month prior to randomization were excluded from the study. Subjects with uncontrolled diabetes mellitus and/or hypertension, subjects with known tuberculosis, HIV, ischemic heart disease, cancer, kidney failure, subjects with significant abnormal laboratory parameters, subjects with known hypersensitivity to any of the ingredients of AQUATURM^®^ were excluded from the study. Other conditions, which in the opinion of the investigators, could have made subject unsuitable for enrolment or could have interfered with his participation in, and completion of the protocol were also excluded from the study.

#### Laboratory Investigations

All the subjects in subset-I of the study were asked to undergo lab testing i.e. Creatinine Kinase (CK) and C reactive protein (CRP) on day 1 (0 hour), day 1 (6 hour), day 2 (24 hour) and day 3 (48 hour). For subset-II, these investigations were performed on day 1, day 2.5 baseline, 24 hours after baseline and 48 hours after baseline.

#### Sample size

With 20 participants in each of the two subsets, the sample size was considered sufficient for analysis with 80% power, Type I error to be 5% and Type II error to be 20%. A total of 40 participants were randomized in the study, 20 in each of the two study subsets.

#### Randomization

Block randomization with centre as the stratum was performed. Subjects meeting eligibility criteria were randomized 1:1 to one of the two groups.

#### Intervention Details

In subset-I, subjects were asked to consume given product in a dose of 1 capsule (250 mg) at 0-hour and one capsule two times a day for 2 days after meals. In subset-II, subjects were asked to consume given product in a dose of 1 capsule (250 mg) twice daily after meals for 5 days.

#### Primary and secondary outcome measures

Primary outcome measures were comparative change in muscle pain on VAS and muscle tenderness between two groups.

Secondary outcome measures were comparative change in muscle swelling, jump performance, muscle damage and inflammation by assessing creatinine kinase (CK) and C reactive protein (CRP), requirement of Tab Paracetamol / NSAIDs as rescue medicine, global assessment for overall change by investigator and subject on CGI Scale, assessment of tolerability of study product by investigator and subject and assessment of adverse events.

#### Assessment of Safety

Safety of the study products was assessed by evaluating adverse events, overall safety and tolerability of the products by the physician and subject on global assessment scale. All adverse events data were listed per subject including severity grading, relationship with investigational product and relationship of the adverse event to other causality, action taken and outcome of the adverse event.

#### Study methodology

A written informed consent was obtained from subjects for their participation in the study before screening. Subjects were recruited if they met all the inclusion criteria. Subject’s demographic details and medical history were noted. Subjects general and systemic examinations, including vitals were performed.

In both subsets, subject’s muscle pain (at Gluteal, Quadriceps, single-leg squat, while walking downstairs, while passive stretch of the quadriceps, passive stretch of the gluteal and while single leg vertical jump) was assessed on Visual Analogue Scale (VAS). Muscle tenderness was graded as 0= none, 1=mild, 2 = moderate and 3= severe. Tenderness was assessed at 4 standardized points. The quadriceps points were marked on the anterior thigh along a line drawn from the anterior superior iliac spine to the superior pole of the patella. One point was at the midpoint of this line (mid belly rectus femoris) and the other at 5 cm above the superior pole of the patella (musculotendinous junction). Muscle swelling was evaluated using non-stretch anthropometric measuring tape at three standardized points on the upper leg. The readings were noted in centimeters. The first point was 5 cm above the superior pole of the patella, the second at the midpoint of a line drawn between the proximal pole of the patella and anterior superior iliac spine and the third at the sub gluteal line. Single-leg vertical squat jump was used to assess the effect of study medications on quadriceps and gluteal function. The jump and reach method were used to assess vertical jump performance. The maximum height obtained from three jumps was recorded. Vertical jump performance was an index of muscle power of the lower limbs. Muscle damage and inflammation were evaluated by assessing serum creatinine kinase (CK) and C reactive protein (CRP). Use of Tab Paracetamol / NSAIDs as rescue medicine was recorded. In subset-I, at 6 hours and in subset-II at day 2.5, subjects were asked to perform a muscle damaging protocol exercise. Subjects were asked to perform seven sets of ten eccentric single-leg press repetitions on a leg press machine. One repetition maximum (1RM) weight was determined for each participant. 120 % of the one repetition maximum was calculated on day one of the study and used for the weight lowered eccentrically. Each participant performed 5 sets of 10 repetitions at 120 % 1RM and 2 sets of 10 repetitions at 100 % 1RM. 1RM was performed on each leg individually and each participant completed all 7 sets. During each eccentric contraction the load was resisted with the allocated leg from full knee extension to 90° angle of knee flexion with the eccentric contraction lasting for 3–5 s duration. After each eccentric contraction the load was raised by subject, using both the legs concentrically. Participants had taken a 3 min rest between sets.

At the end of study, subject’s global evaluation and investigator’s global evaluation for overall change was done on last follow up visit. Tolerability of the study products was assessed by investigator and by subject. All the subjects were closely monitored for any adverse events starting from baseline visit till the end of the study.

#### Plan for statistical analysis

The data were analysed for central tendencies (mean, median), range, standard error and standard deviation. Data were tabulated and graphically shown using standard format and MS Excel. Statistical tests were carried out to compare study groups as per the distribution (normality) Student’s T test (normative), Mann-Whitney statistic (non-parametric), Chi-square statistic (categorical), ANOVA. The level of significance at *p* < 0.05 (two sided) was considered significant. Both intent-to-treat and per protocol completer analysis were performed when appropriate. Standard statistical software programs were used (GraphPad InStat Version 3.6).

## Results

### Subset-I

#### Baseline Demography

A total of 20 subjects completed the study. The details are presented in the CONSORT chart. No significant difference between the two groups with respect to baseline demography was observed. The details are presented in Table 1.

**Table 1:** Baseline Demography.

| Demography | AQUATURM<sup>®</sup> | Placebo |
| --- | --- | --- |
| Subjects (male) | 10 | 10 |
| Age in years | 22.1±4.31 | 20.2±2.1 |
| Height in Meter | 1.72±0.05 | 1.74±0.05 |
| Weight in Kg | 70.67±9.90 | 76.03±10.49 |
| BMI | 24±3.03 | 24.9±2.97 |

#### Primary Outcomes

##### Assessment of changes in muscle pain on VAS between two groups

Statistically significant reduction in muscle pain (at Gluteal, Quadriceps, single-leg squat, walking downstairs, passive stretch of the quadriceps, passive stretch of the gluteal and single leg vertical jump) was observed in the AQUATURM^®^ group at 24 hours, and 48 hours compared to the placebo group. The details are presented in table 2.

**Table 2:**
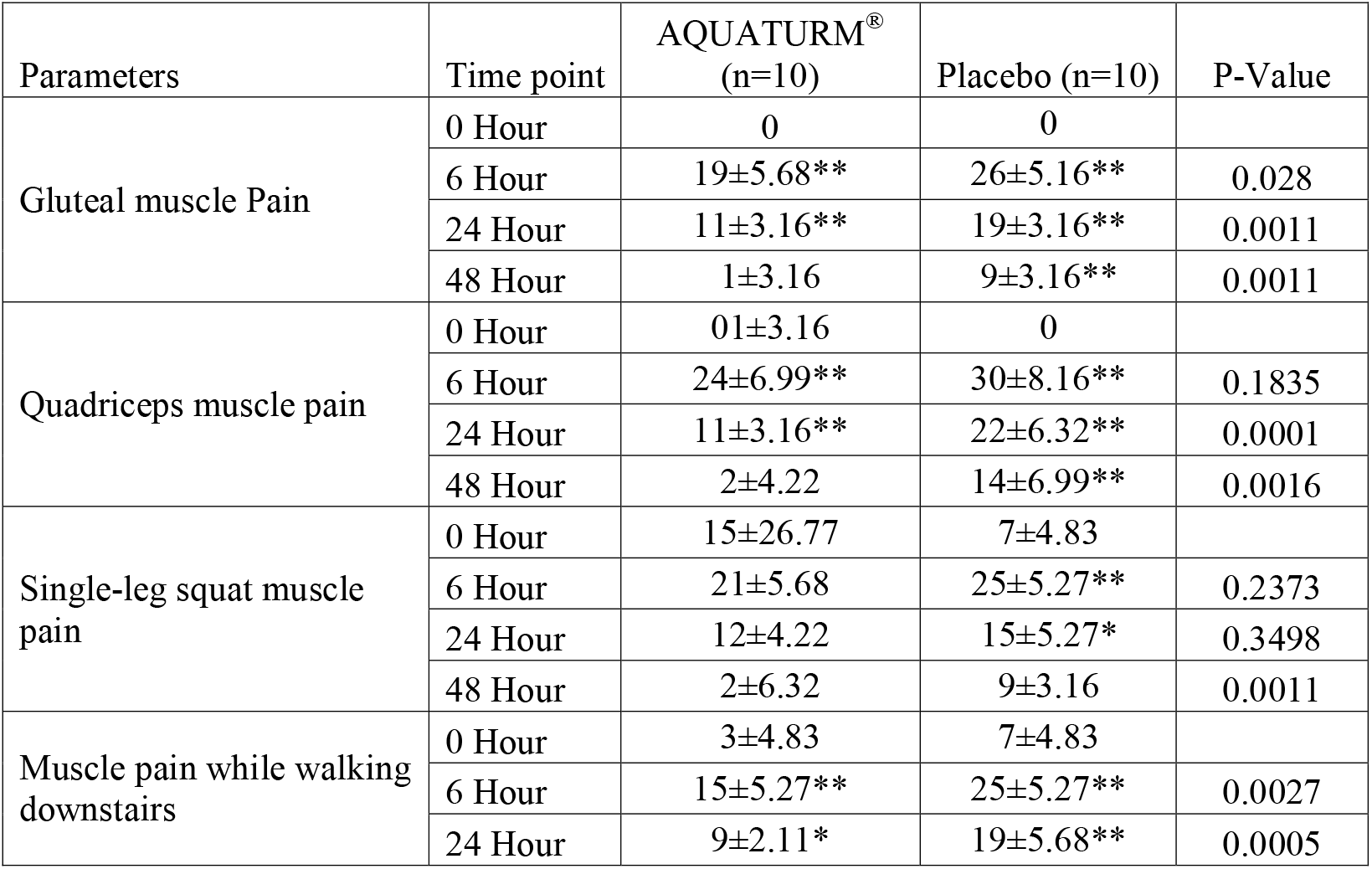
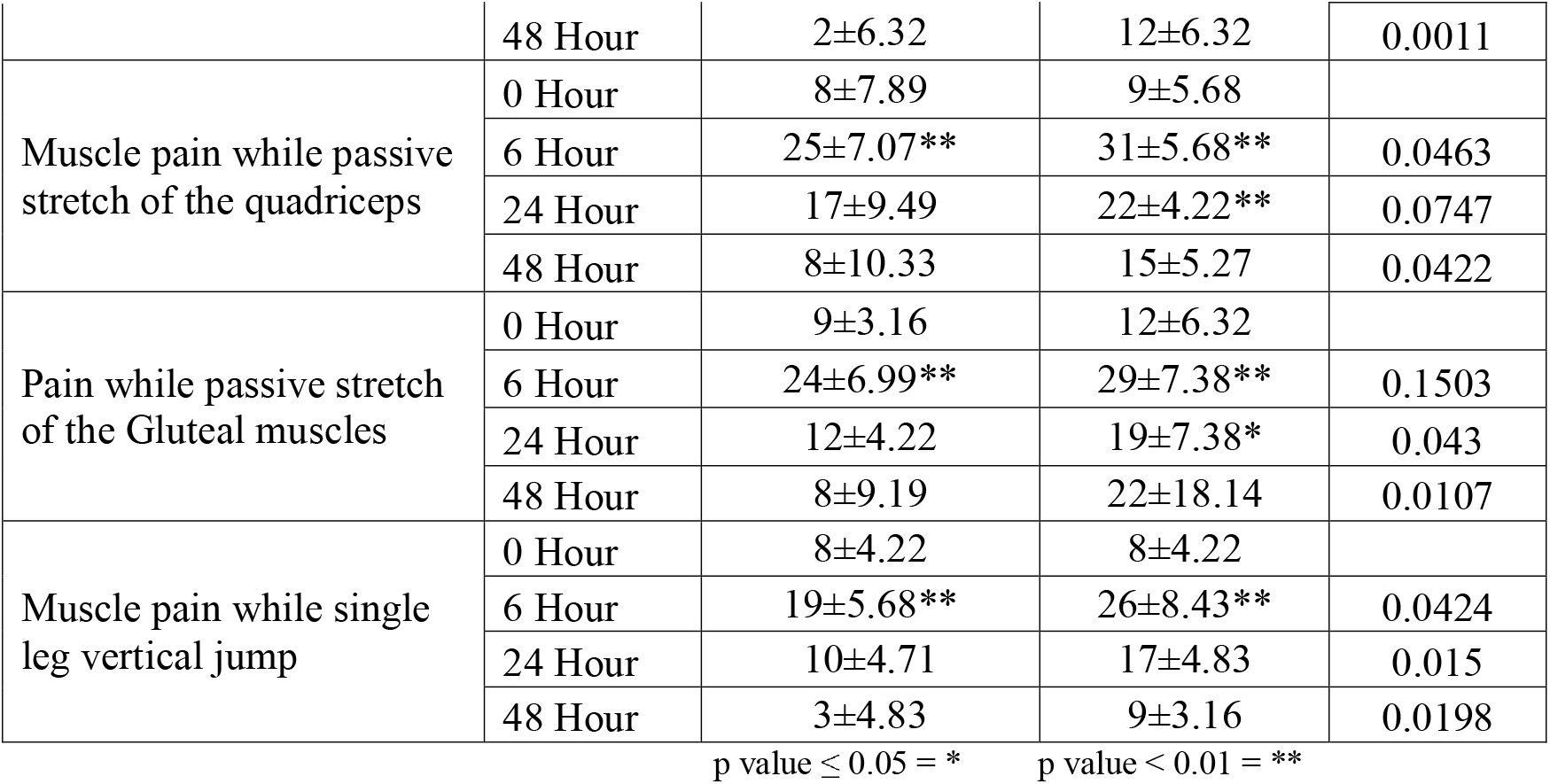
Assessment of changes in muscle pain on VAS between two groups:

##### Assessment of change in muscle tenderness between two groups

Statistically significant reduction in muscle tenderness at mid belly rectus femoris and musculotendinous junction was observed in the AQUATURM^®^ group at 48 hours as compared to the Placebo group. The details are presented in figure 1 and 2.

**Figure 1:**
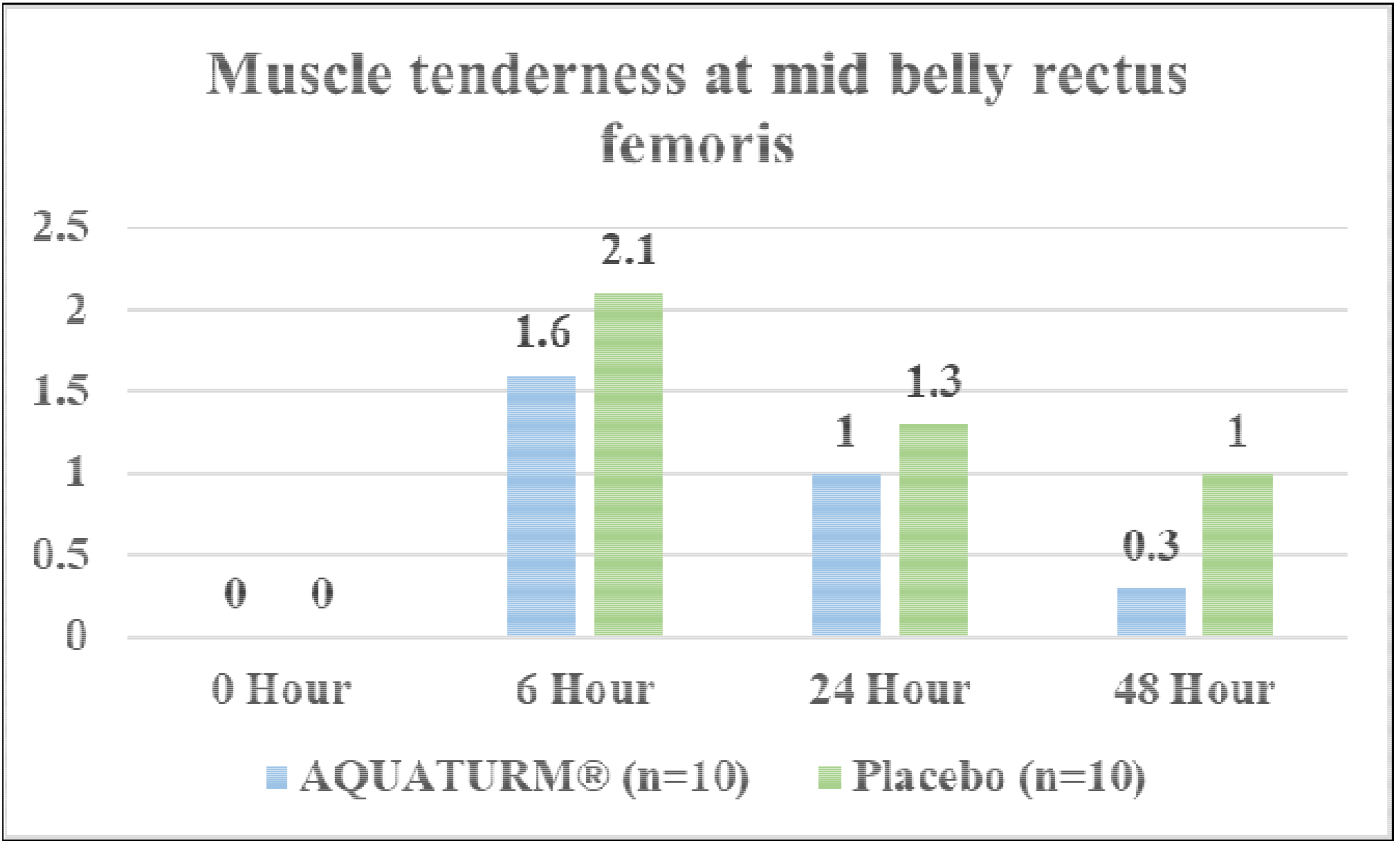
Assessment of muscle tenderness at mid belly rectus femoris between the two groups:

**Figure 2:**
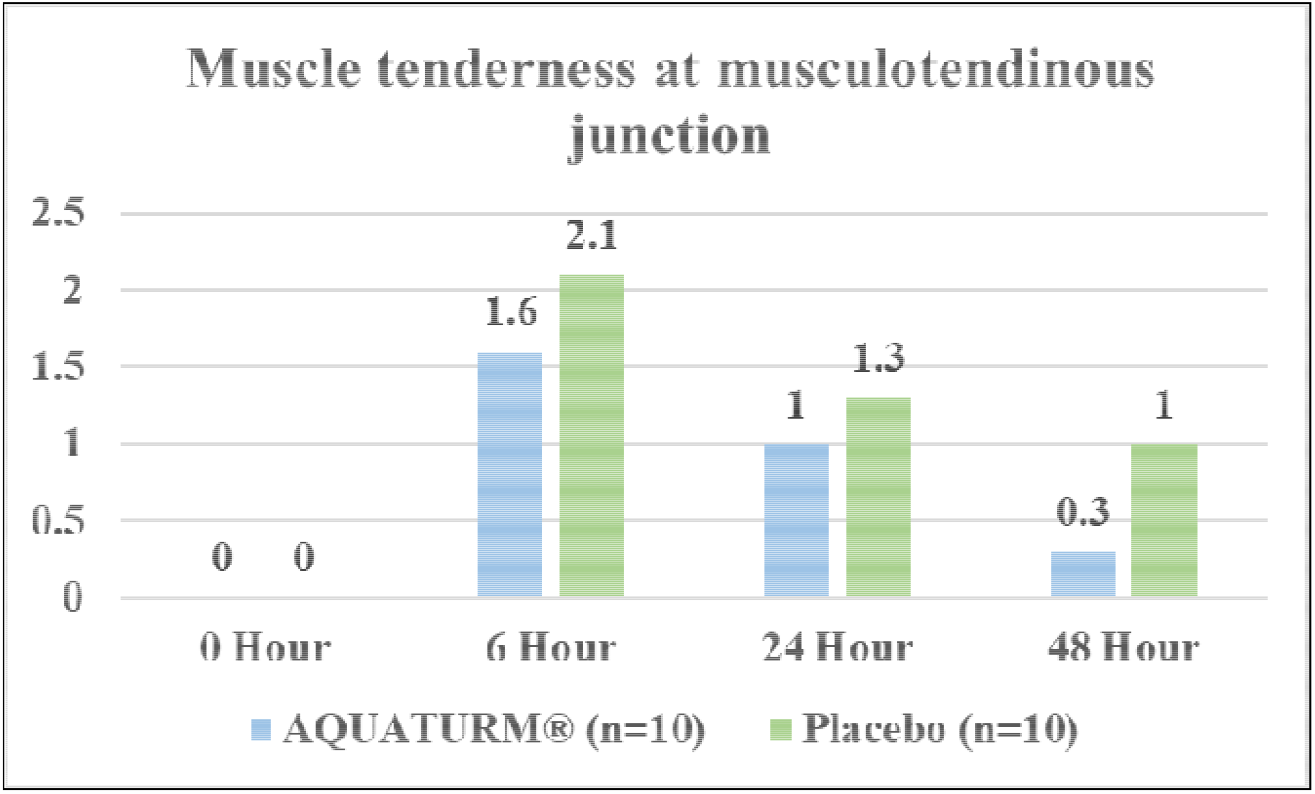
Assessment of muscle tenderness at musculotendinous junction between the two groups:

#### Secondary Outcomes

##### Assessment of changes in muscle swelling between two groups

Statistically significant reduction in muscle swelling at midpoint of a line drawn between the proximal pole of the patella and anterior superior iliac spine was observed in the AQUATURM^®^ group at 48 hours compared to the placebo group. Between group analysis showed no significant difference in reduction of muscle swelling at 5 cm above the superior pole of the patella and sub gluteal line. The details are presented in table 3.

**Table 3:** Assessment of changes in muscle swelling between two groups:

| Parameters | Time Points | AQUATUM® (n=10) | Placebo (n=10) | P-Value |
| --- | --- | --- | --- | --- |
| Muscle Swelling at 5 cm above the superior pole of the patella(cm) | 0 Hour | 39.70 ±2.50 | 41.70 ±3.97 |  |
|  | 6 Hour | 40.70 ±2.50** | 43 ±4.40** | 0.1675 |
|  | 24 Hour | 39.90 ±2.23 | 42.70 ±3.97** | 0.1791 |
|  | 48 Hour | 39.60 ±2.41 | 42.40 ±4.27* | 0.2049 |
| Muscle Swelling at midpoint of a line drawn between the proximal pole of the patella and anterior superior iliac spine(cm) | 0 Hour | 48.80±3.91 | 50.30±2.71 |  |
|  | 6 Hour | 50.80±3.97** | 51.90±2.77** | 0.4812 |
|  | 24 Hour | 49.70±3.80** | 51.50±2.51** | 0.2273 |
|  | 48 Hour | 48.50±3.24 | 51.70±2.75** | 0.0285 |
| Muscle Swelling at sub gluteal line (cm) | 0 Hour | 94.10±7.06 | 99.40±7.85 |  |
|  | 6 Hour | 96.10±7.06** | 101.30±7.97** | 0.14 |
|  | 24 Hour | 94.70±6.82** | 100.60±7.79** | 0.0883 |
|  | 48 Hour | 93.90±6.74 | 100.40±8.04** | 0.0658 |

p value ≤ 0.05 = \* p value < 0.01 = \*\*

##### Assessment of change in jump performance between two groups

No significant difference in the average of (first, second and third) single-leg vertical squat jump was observed between the two groups. The details are presented in table 4.

**Table 4:** Assessment of change in jump performance between two groups:

| Parameter | Time | AQUA-TURM<sup>®</sup><br>(n=10) | Placebo<br>(n=10) | P-Value |
| --- | --- | --- | --- | --- |
| Average of (First, Second and Third) Single-leg vertical squat jump (cm) | 0 Hour | 10.37 ±0.74 | 9.73 ±0.81 |  |
|  | 6 Hour | 9.47 ±0.71** | 9.68 ±1.39 | 0.3034 |
|  | 24 Hour | 10.33 ±0.67 | 10.13±1.20 | 0.8866 |
|  | 48 Hour | 10.73 ±1.03 | 10.3 ±0.86 | 0.1601 |

p value ≤ 0.05 = \* p value < 0.01 = \*\*

##### Assessment of change in muscle damage and inflammation by assessing Creatinine Kinase (CK) and C reactive protein (CRP) between two groups

There was no significant change from baseline to end of study in CPK and CRP levels in both AQUATURM^®^ and Placebo Groups. Between group analysis showed no significant difference. The details are presented in table 5.

**Table 5:** Assessment of change in muscle damage and inflammation by assessing Creatinine Kinase (CK) and C reactive protein (CRP) between two groups:

| Lab investigations | AQUATURM® |  | Placebo |  | P Value (Between) |
| --- | --- | --- | --- | --- | --- |
|  | Pre | Post | Pre | Post |  |
| CPK | 390.81±418.44 | 301.76±182.19 | 281.92±101.30 | 304.58±268.67 | 0.6334 |
|  |  | 0.2871 |  | 0.7422 |  |
| CRP | 0.21±0.12 | 0.24±0.23 | 0.24±0.11 | 0.23±0.07 | 0.2358 |
|  |  | 0.4922 |  | 0.1339 |  |

##### Assessment of Rescue Medication between two groups

None of the subjects from both the groups used Tab Paracetamol / NSAIDs as rescue medication during the study period.

##### Assessment of changes in global assessment for overall change by Investigator and Subject on CGI Scale between two groups

In AQUATURM^®^ group, as per investigator and subject’s assessment, 80% subjects reported very much improved, whereas in placebo group, as per investigators and subject’s assessment, 60% subjects reported minimum worsening in their condition. When compared between the two groups, the difference was statistically significant. The details are presented in table 6.

**Table 6:** Assessment of changes in global assessment for overall change by Investigator and Subjects on CGI Scale between two groups:

| Parameters of CGI Scale | By Investigator |  | By Subject |  |
| --- | --- | --- | --- | --- |
|  | AQUATURM® | Placebo | AQUATURM® | Placebo |
| Very Much Improved=1 | 8 (80%) | 1 (10%) | 8 (80%) | 1 (10%) |
| Much Improved=2 | 1 (10%) | 0 (0%) | 1 (10%) | 0 (0%) |
| Minimally Improved= 3 | 1(10%) | 0 (0%) | 1(10%) | 0 (0%) |
| No Change=4 | 0 (0%) | 3 (30%) | 0 (0%) | 3 (30%) |
| Minimally Worse=5 | 0 (0%) | 6 (60%) | 0 (0%) | 6 (60%) |
| Total | 10 (100%) | 10 (100%) | 10 (100%) | 10 (100%) |

#### Safety Parameters

##### Assessment of tolerability of study products and adverse events

Both the study products were very well tolerated by all the subjects of both the groups. No adverse event was reported in AQUATURM^®^ group. In Placebo group, 01 adverse event (AE) i.e., headache was reported. No treatment or interruption of the study product was required to resolve the event. As per investigator’s assessment AE was possibly related to the study products.

##### Assessment of vitals

There was no significant difference from baseline to all the follow up visits in all the vital parameters like pulse rate, respiration rate, body temperature and blood pressure in both the groups. The levels were within normal range at all visits in both the study groups.

### Subset-II

#### Base Line Demography

A total of 20 subjects completed the study. The details are presented in the CONSORT chart. No significant difference between the two groups with respect to baseline demography was observed. The details are presented in Table 7.

**Table 7:** Baseline Demography.

| **Demography** | **AQUATUM<sup>®</sup>** | **Placebo** |
| --- | --- | --- |
| Male | 10 | 10 |
| Age | 22.10±3.87 | 23.40±4.65 |
| Height in Meter | 1.73±0.06 | 1.699±0.054 |
| Weight in Kg | 70.92±11.31 | 67.06±8.24 |
| BMI | 23.7±2.9 | 23.2±2.82 |

#### Primary Outcomes

##### Assessment of changes in muscle pain on VAS between two groups

Statistically significant reduction in muscle pain (at Quadriceps, single-leg squat) was observed in the AQUATURM^®^ group at 24 hours after baseline compared to the placebo group. Statistically significant reduction in muscle pain (at Gluteal, while walking downstairs, while passive stretch of the quadriceps, passive stretch of the gluteal and while single leg vertical jump) was observed in the AQUATURM^®^ group at 24 hours and 48 hours after baseline compared to the placebo group. The details are presented in table 8.

**Table 8:** Assessment of changes in muscle pain on VAS between two groups:

| Parameters | Time Points | AQUATURM<sup>®</sup><br>(n=10) | Placebo<br>(n=10) | P Value |
| --- | --- | --- | --- | --- |
| Gluteal Muscle Pain | 0 Hour | 0 | 0 |  |
|  | Baseline visit<br>(BV) (2.5 Days) | 21 ±7.38** | 25 ±5.27** | 0.3089 |
|  | 24 Hour after BV | 11 ±5.68** | 19 ±3.16** | 0.0049 |
|  | 48 Hour after BV | 1 ±3.16 | 11 ±3.16** | 0.0001 |
| Quadriceps Muscle pain | 0 Hour | 0 | 0 |  |
|  | BV (2.5 Days) | 23 ±4.83** | 27 ±4.83** | 0.1789 |
|  | 24 Hour after BV | 11 ±5.68** | 19 ±3.16** | 0.0049 |
|  | 48 Hour after BV | 1 ±3.16 | 11 ±3.16** | 0.0001 |
| Single-leg squat muscle pain | 0 Hour | 4 ±5.16 | 3 ±4.83 |  |
|  | BV (2.5 Days) | 26 ±5.16** | 27 ±4.83** | >0.9999 |
|  | 24 Hour after BV | 13 ±6.75 | 20±0** | 0.0108 |
|  | 48 Hour after BV | 1 ±3.16 | 13 ±4.83* | <0.0001 |
| Muscle pain while walking<br>downstairs | 0 Hour | 4 ±5.16 | 3 ±4.83 |  |
|  | BV (2.5 Days) | 26 ±5.16** | 29 ±5.67** | 0.4195 |
|  | 24 Hour after BV | 13 ±6.75 | 20±0** | 0.0108 |
|  | 48 Hour after BV | 2 ±4.22 | 14 ±5.16** | 0.0003 |
| Muscle pain while passive<br>stretch of the quadriceps | 0 Hour | 9 ±3.16 | 8 ±4.21 |  |
|  | BV (2.5 Days) | 35 ±5.27** | 37 ±4.83** | 0.6499 |
|  | 24 Hour after BV | 20 ±4.71** | 27 ±4.83** | 0.015 |
|  | 48 Hour after BV | 10 ±4.71 | 20 ±4.71** | 0.0009 |
| Pain while passive stretch of the gluteal muscles | 0 Hour | 9 ± 3.16 | 8 ± 4.21 | |
| | BV (2.5 Days) | 35 ± 5.27** | 37 ± 4.83** | 0.6499 |
| | 24 Hour after BV | 19 ± 5.68** | 27 ± 4.83** | 0.011 |
| | 48 Hour after BV | 9 ± 5.68 | 20 ± 4.71** | 0.0008 |
| Muscle pain while single leg vertical jump | 0 Hour | 6 ± 5.16 | 6 ± 5.16 | |
| | BV (2.5 Days) | 34 ± 5.16** | 34 ± 5.16** | >0.9999 |
| | 24 Hour after BV | 16 ± 6.99* | 24 ± 5.16** | 0.0186 |
| | 48 Hour after BV | 4 ± 5.16 | 18 ± 6.32** | 0.0004 |

p value ≤ 0.05 = \*
p value < 0.01 = \*\*

##### Assessment of change in muscle tenderness between two groups

Statistically significant reduction in muscle tenderness at mid belly rectus femoris and musculotendinous junction was observed in the AQUATURM^®^ group at 48 hours after baseline compared to the Placebo group. The details are presented in figure 3 and 4.

**Figure 3:**
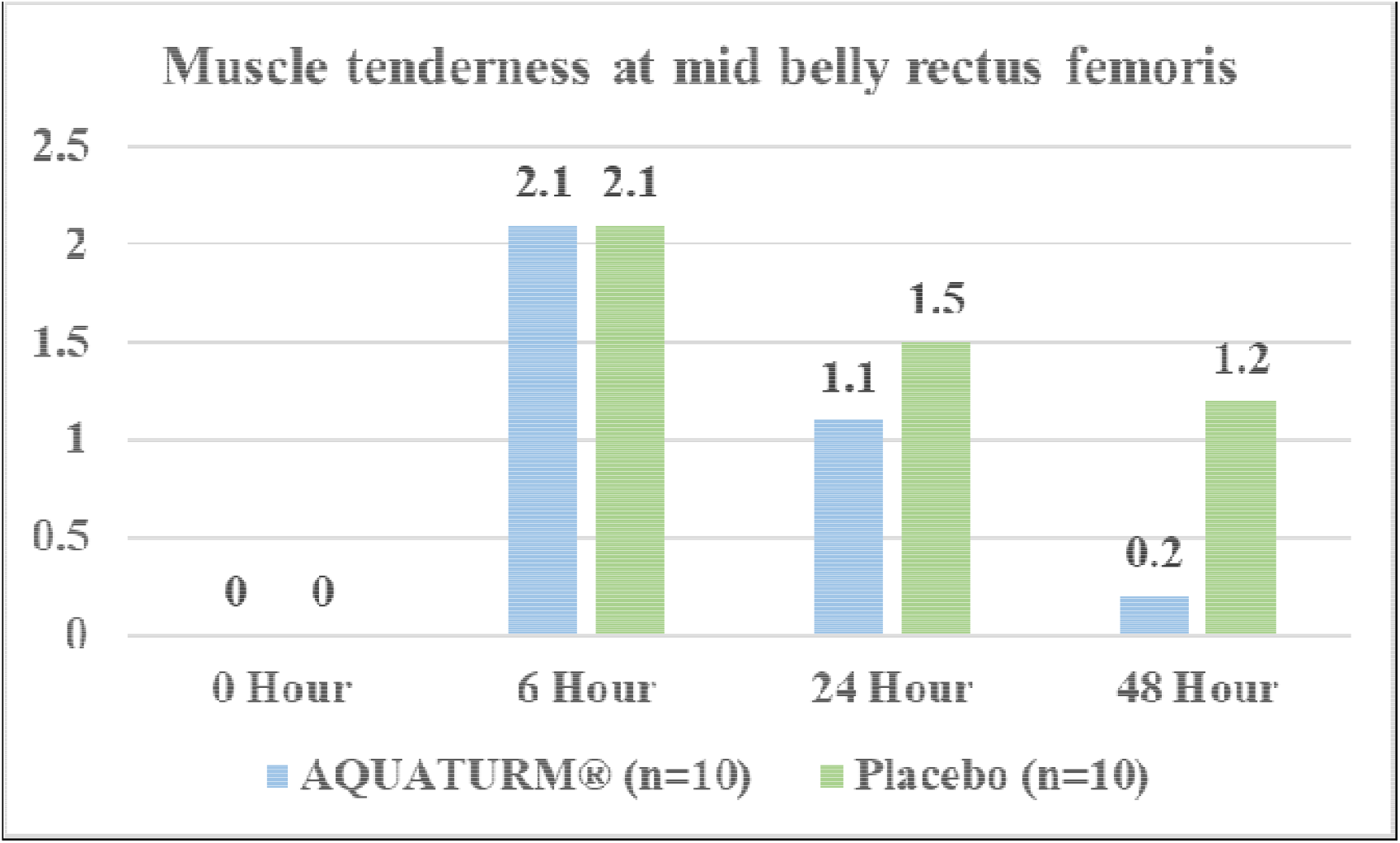
Muscle tenderness at mid belly rectus femoris.

**Figure 4:**
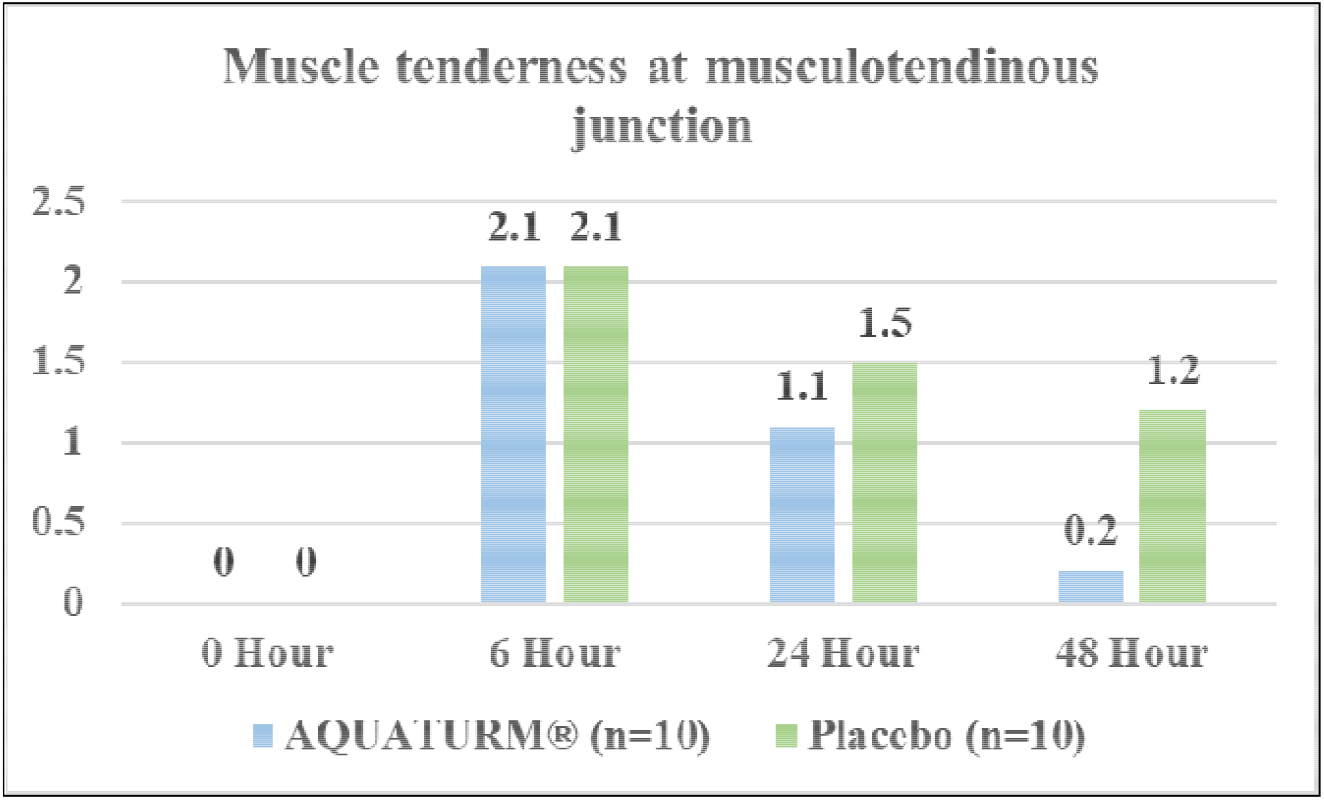
Muscle tenderness at musculotendinous junction.

#### Secondary Outcomes

##### Assessment of changes in muscle swelling between two groups

No statistically significant difference in the reduction of muscle swelling at 5 cm above the superior pole of the patella(cm), midpoint of a line drawn between the proximal pole of the patella and anterior superior iliac spine and sub gluteal line was observed between the two groups. The details are presented in table 9.

**Table 9:** Assessment of changes in muscle swelling between two groups:

| Parameters | Time | AQUATURM®<br>(n=10) | Placebo<br>(n=10) | P<br>Value |
| --- | --- | --- | --- | --- |
| Muscle Swelling at 5 cm<br>above the superior pole of<br>the patella(cm) | 0 Hour | 40 ±2.40 | 38 ±2.10 |  |
|  | BV (2.5 Days) | 41 ±2.40** | 39.80 ±2.10** | 0.2497 |
|  | 24 Hour after BV | 40.10 ±2.42 | 39.60 ±2.22** | 0.6364 |
|  | 48 Hour after BV | 40.10 ±2.42 | 39.20 ±2.62 | 0.4353 |
| Muscle Swelling at midpoint<br>of a line drawn between the<br>proximal pole of the patella<br>and anterior superior iliac<br>spine(cm) | 0 Hour | 48 ±3.23 | 47.50 ±2.88 |  |
|  | BV (2.5 Days) | 49.60 ±3.03** | 49.30 ±3.06** | 0.9263 |
|  | 24 Hour after BV | 48.60 ±3.03** | 48.50 ±3.10 | 0.4895 |
|  | 48 Hour after BV | 48 ±3.23 | 47.60 ±3.31 | 0.7875 |
| Muscle Swelling at sub<br>gluteal line (cm) | 0 Hour | 93.30 ±7.44 | 92.40 ±5.25 |  |
|  | BV (2.5 Days) | 95.20 ±7.35** | 94.40 ±5.23** | 0.7823 |
| | 24 Hour after BV | 94.20 ± 7.35** | 93.50 ± 5.25** | 0.8092 |
| | 48 Hour after BV | 93.30 ± 7.44 | 92.80 ± 5.27* | 0.8642 |

p value ≤ 0.05 = \* p value $<$ 0.01 = \*\*

##### Assessment of change in jump performance between two groups

Statistically significant increase in average single-leg vertical squat jump was reported in the AQUATURM^®^ group at 24 hour and 48 hours after baseline visit as compared to the Placebo group. The details are presented in table 10.

**Table 10:**
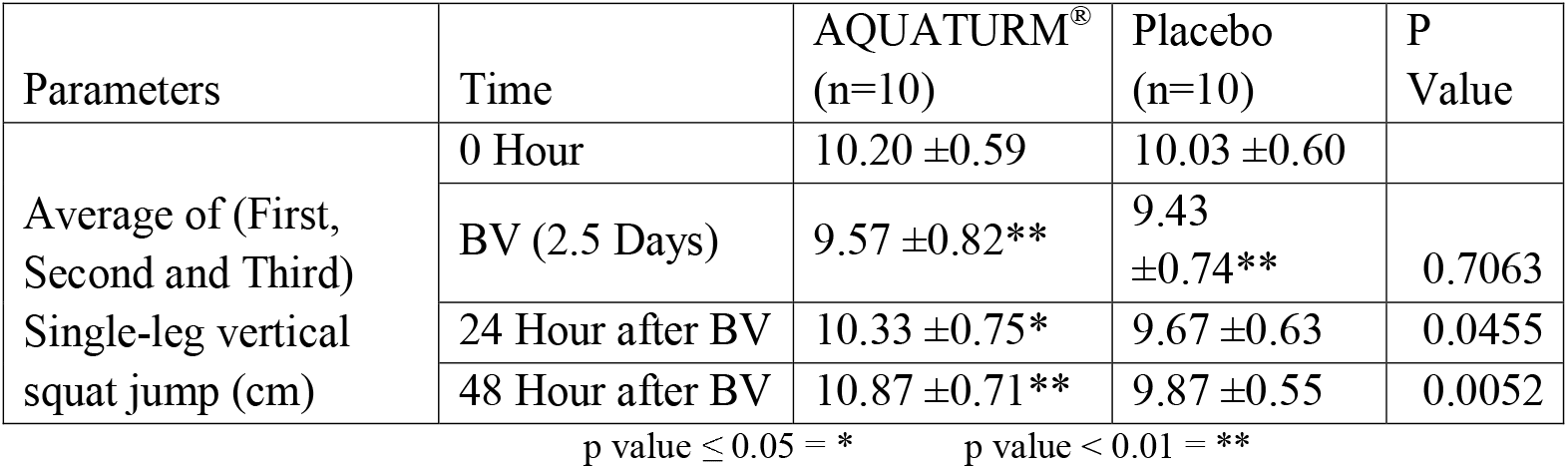
Assessment of change in jump performance between two groups:

##### Assessment of change in muscle damage and inflammation by assessing Creatinine Kinase (CK) and C reactive protein (CRP) between two groups

There was no significant change from baseline to end of study in CPK and CRP levels in both AQUATURM^®^ and Placebo Group. Between group analysis showed no significant difference. The details are presented in table 11.

**Table 11.** Assessment of change in muscle damage and inflammation by assessing Creatinine Kinase (CK) and C reactive protein (CRP) between two groups:

|  |  | AQUATUM<sup>®</sup> |  | Placebo |  | P Value<br>(Between) |
| --- | --- | --- | --- | --- | --- | --- |
|  |  | Pre | Post | Pre | Post |  |
| CPK | Mean ± SD | 656.62 ± 867.19 | 310.94 ± 207.40 | 300.12 ± 190.46 | 291.42 ± 261.66 | 0.4813 |
|  | P Value |  | 0.2324 |  | 0.5566 |  |
| CRP | Mean ± SD | 0.17 ± 0.05 | 0.31 ± 0.20 | 0.20 ± 0.11 | 0.18 ± 0.05 | 0.0734 |
|  | P Value |  | 0.0488 |  | 0.8457 |  |

##### Assessment of Rescue Medication between two groups

None of the subjects from both the groups used Tab Paracetamol / NSAIDs as rescue medication during the study period.

##### Assessment of changes in global assessment for overall change by Investigator and Subject on CGI Scale between two groups

In AQUATURM^®^ group, as per investigators and subject’s assessment, 70% and 80% subjects reported very much improvement respectively. In Placebo group, as per investigators and subject’s assessment, 40% and 30% subjects reported minimum worsening, respectively. When compared between the two groups, the difference was statistically significant. The details are presented in table 12.

**Table 12:** Assessment of changes in global assessment for overall change by Investigator and Subject on CGI Scale between two groups:

|  | By Investigator |  | By Subject |  |
| --- | --- | --- | --- | --- |
|  | AQUATURM<sup>®</sup> | Placebo | AQUATURM<sup>®</sup> | Placebo |
| Very Much Improved=1 | 7 (70%) | 0 (0%) | 8 (80%) | 0 (0%) |
| Much Improved=2 | 2 (20%) | 0 (0%) | 1 (10%) | 0 (0%) |
| Minimally Improved= 3 | 1 (10%) | 5 (50%) | 1 (10%) | 5 (50%) |
| No Change=4 | 0 (0%) | 1 (10%) | 0 (0%) | 2 (20%) |
| Minimally Worse=5 | 0 (0%) | 4 (40%) | 0 (0%) | 3 (30%) |
| Total | 10 (100%) | 10 (100%) | 10 (100%) | 10 (100%) |

#### Safety Parameters

##### Assessment of tolerability of study products and assessment of adverse events

Both the study products were very well tolerated by all the subjects of both the groups. No adverse event was reported in any of the subjects of both the study groups.

##### Assessment of vitals

There was no significant difference from baseline to all the follow up visits in all the vital parameters like pulse rate, respiration rate, body temperature and blood pressure in both the groups. The levels were within normal range at all visits in both the study groups.

## Discussion

The present study was conducted to demonstrate efficacy and safety of AQUATURM® Capsule that contains 250 mg water soluble turmeric extract standardized to not less than 30% total Curcuminoids in subjects suffering from Delayed onset muscle soreness (DOMS). The study was conducted in two subsets in order to evaluate efficacy and safety of consumption of AQUATURM® Capsule for 2 days (subset-I) and 5 days (subset-II) in subjects suffering from DOMS.

In both the subsets, no significant difference between the two groups with respect to baseline demography was observed. The results of the present study demonstrated a statistically significant reduction in muscle pain (at gluteal, quadriceps, single-leg squat, while walking downstairs, while passive stretch of the quadriceps, passive stretch of the gluteal and while single leg vertical jump) post eccentric exercise in AQUATURM® Capsule group compared to placebo group in both the subsets of the study. Statistically significant reduction in muscle swelling (midpoint of a line drawn between the proximal pole of the patella) post eccentric exercise was observed in AQUATURM® Capsule group compared to placebo group in the subset-I of the study. Also, a statistically significant reduction in muscle tenderness (at mid belly rectus femoris and musculotendinous junction) post eccentric exercise was observed in AQUATURM® Capsule group compared to placebo group in both the subsets of the study. Previous studies have demonstrated alleviation of symptoms of DOMS with the use of curcumin supplementation.^11^

The anti-inflammatory effect of AQUATURM® Capsule to relieve muscle pain post eccentric exercise may be related to the desensitization or the inhibition of a series of transient receptor potential ion channels involved in the generation of painful stimuli like TRPV1 and TRPA1 ^15, 16^ Several studies have confirmed that curcumin down-regulates the expression of several pro-inflammatory cytokines involved in proteolysis and muscle inflammation^17^ by suppressing NF— κB signalling. ^18, 19^ Curcumin can reduce the expression of the inflammatory markers intercellular adhesion molecule 1 (ICAM-1) and vascular cell adhesion molecule 1 (VCAM- 1). Curcumin attenuates the activity of 5-lipoxygenase (5-LOX) and cyclooxygenase-2 (COX- 2), which are enzymes in the leukotriene-producing metabolic pathway.^14^ These mechanisms could explain the significant effect of AQUATURM® Capsule in relieving muscle tenderness and swelling post exercise.

Statistically significant increase in average single-leg vertical squat jump was reported in the AQUATURM^®^ group at 24 hour and 48 hours after baseline visit as compared to the Placebo group suggesting improvement in muscle power of the lower limbs. Since AQUATURM® Capsule effectively relieved muscle pain, tenderness and swelling post exercise that could have led to improvement in the muscle power of lower limbs.

Post supplementation, no significant difference in CPK and CRP levels were observed between the groups in both the subsets of the study, however better reduction was observed in AQUATURM^®^ group. Also, none of the subjects from either group used Tab Paracetamol / NSAIDs as rescue medication during the study period.

AQUATURM® Capsule was well tolerated by the subjects as there were no side effects reported during the study. Post study, no significant change in any of the vital parameters such as pulse rate, respiratory rate, temperature, systolic and diastolic blood pressure was observed. This suggests safety of AQUATURM® Capsule.

## Conclusion

It can be concluded from the results of the study that supplementation of AQUATURM® Capsule prior to and following heavy eccentric exercise in healthy men results in relieving muscle pain, tenderness and swelling caused due to DOMS. Also, AQUATURM® Capsule helps to improve muscle power post strenuous exercise. AQUATURM® Capsule was well tolerated by the subjects as there was no side effects reported during the study. Additional studies on DOMS with AQUATURM® are encouraged.

## Data Availability

All data produced in the present work are contained in the manuscript.

## Authors’ Contributions

All authors are responsible for the content and writing of the paper. All authors have critically contributed to the conceptualization, data collection, analysis, and writing of the paper.

## Acknowledgments

Authors greatly acknowledge the invaluable collaborative efforts of the study participants. Authors express their gratitude to Lodaat Pharma for sponsoring this study. Authors also thank team at Target Institute and Medical Education and Research, Mumbai, India and Lodaat Pharma, Illinois, US for their priceless support.

---

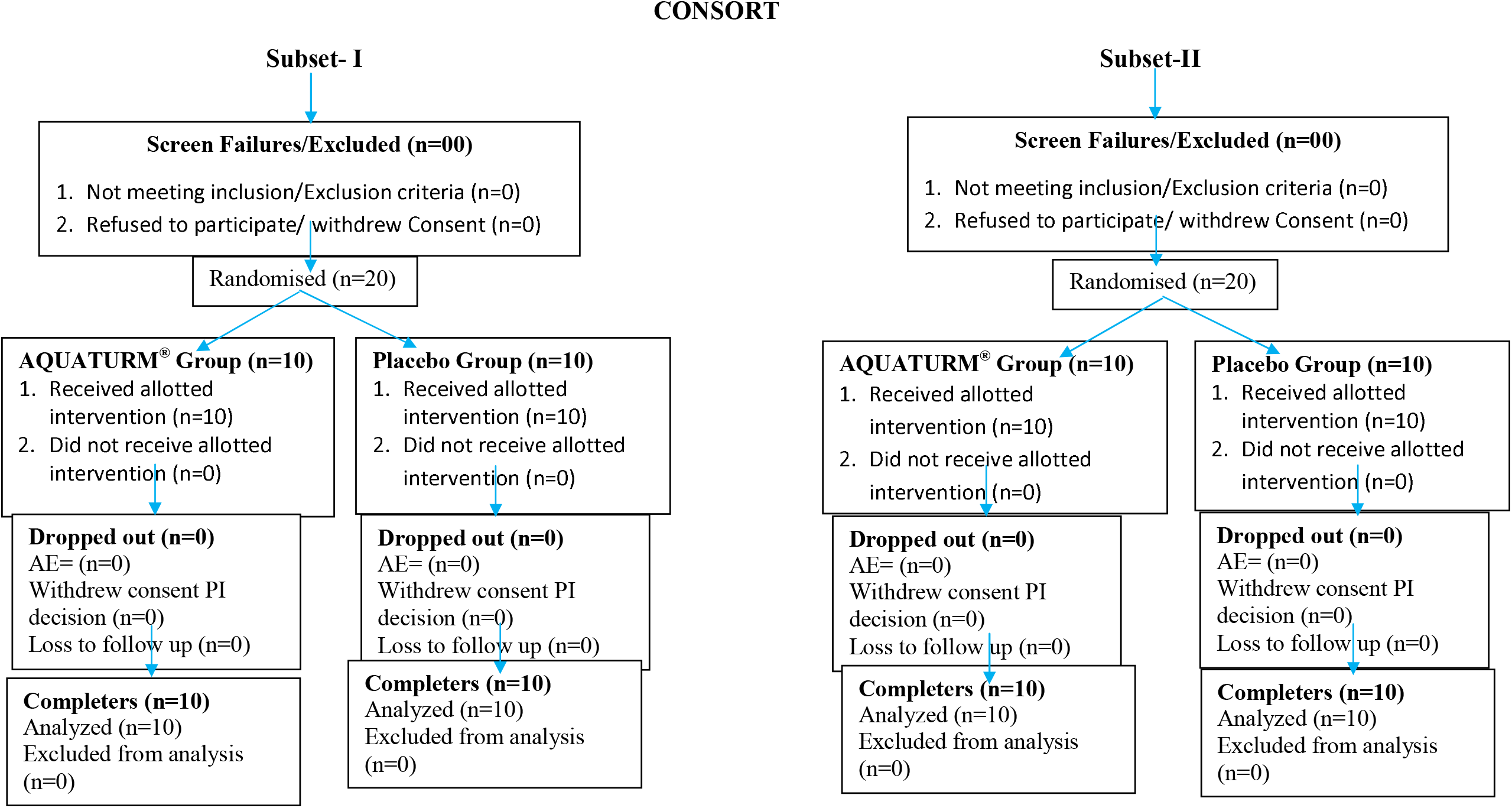

## References

1. PC Zondi, DC Janse van Rensburg, CC Grant, A Jansen van Rensburg. Delayed onset muscle soreness: No pain, no gain? The truth behind this adage. South African Family Practice 2015; 57(57):29–33

2. Kazue Mizumura, Toru Taguchi. Delayed onset muscle soreness: Involvement of neurotrophic factors. J Physiol Sci (2016) 66:43–52

3. M.J. Cleak & R.G. Eston. Delayed onset muscle soreness: Mechanisms and management. Journal of Sports Sciences. Volume 10, 1992 - Issue 4, Pages 325–341

4. Jeffrey C. Stay et. al. Pulsed Ultrasound Fails To Diminish Delayed-Onset Muscle Soreness Symptoms. Journal of Athletic Training 1998;33(4):341–346

5. J E Hilbert, G A Sforzo, T Swensen. The effects of massage on delayed onset muscle soreness. Br J Sports Med 2003; 37:72–75

6. Ruth o’connor and Deirdre A. Hurley. The effectiveness of physiotherapeutic Interventions in the management of delayed onset Muscle soreness: a systematic review. Physical Therapy Reviews 2003; 8: 177–195

7. Zubia Veqar and Shagufta Imtiyaz. Vibration Therapy in Management of DOMS. Journal of Clinical and Diagnostic Research. 2014 Jun, Vol-8(6)

8. Karoline Cheung, Patria A. Hume and Linda Maxwell. Delayed Onset Muscle Soreness Treatment Strategies and Performance Factors. Journal of Athletic Training 1998;33(4):341–346

9. Declan A.J. Connolly, Stephen P. Sayers, and Malachy P. Mchugh. Treatment and Prevention of Delayed Onset Muscle Soreness. Journal of Strength and Conditioning Research, 2003, 17(1), 197–208

10. Franchek Drobnic, Joan Riera, Giovanni Appendino, Stefano Togni, Federico Franceschi, Xavier Valle, Antoni Pons5 and Josep Tur. Reduction of delayed onset muscle soreness by a novel curcumin delivery system (Meriva®): a randomised, placebo-controlled trial. Journal of the International Society of Sports Nutrition 2014, 11:31

11. Lesley M. Nicol · David S. Rowlands · Ruth Fazakerly · John Kellett. Curcumin supplementation likely attenuates delayed onset muscle soreness (DOMS). Eur J Appl Physiol (2015) 115:1769–1777

12. Wan-Young Yoon, Kihyuk Lee and Jooyoung Kim. Curcumin supplementation and delayed onset muscle soreness (DOMS): effects, mechanisms, and practical considerations. Phys Act Nutr. 2020 Sep 30; 24(3): 39–43.

13. Asher GN, Spelman K. Clinical utility of curcumin extract. Altern Ther Health Med. 2013;19(2):20–2

14. Wan-Young Yoon, Kihyuk Lee, Jooyoung Kim. Curcumin supplementation and delayed onset muscle soreness (DOMS): effects, mechanisms, and practical considerations. J Exerc Nutrition Biochem. 2018;22(2):007–011

15. Leamy AW, Shukla P, McAlexander MA, Carr MJ, Ghatta S: Curcumin ((E, E)-1,7-bis(4-hydroxy-3-methoxyphenyl)-1,6-heptadiene-3,5-dione) activates and desensitizes the nociceptor ion channel TRPA1. Neurosci Lett 2011, 503:157–162.

16. Yeon KY, Kim SA, Kim YH, Lee MK, Ahn DK, Kim HJ, Kim JS, Jung SJ, Oh SB: Curcumin produces an antihyperalgesic effect via antagonism of TRPV1. J Dent Res 2010, 89:170–174

17. Singh S, Aggarwal BB: Activation of transcription factor NF-kappa B is suppressed by curcumin (diferuloylmethane) [corrected]. J Biol Chem 1995, 270:24995–25000

18. Das L, Vinayak M: Long term effect of curcumin down regulates expression of TNF-alpha and IL-6 via modulation of ETS and NF-kappaB transcription factor in liver of lymphoma bearing mice. Leukemia & lymphoma 2014, In press.

19. Fu Y, Gao R, Cao Y, Guo M, Wei Z, Zhou E, Li Y, Yao M, Yang Z, Zhang N: Curcumin attenuates inflammatory responses by suppressing TLR4-mediated NF-kappaB signaling pathway in lipopolysaccharide-induced mastitis in mice. Int Immunopharmacol 2014, 20(1):54–58

